# Temporal evolution acquisition based arterial spin labeling (TEA-ASL) for accurate arterial blood T_2_ mapping

**DOI:** 10.64898/2026.01.10.26343600

**Authors:** Jiawen Sun, Chenhe Yuan, Junqi Xu, Jiekai Zhu, Ning Wang, Yiling Liu, Qing Wei, Weihuan Fang, Zhensen Chen, Chenyan Wang, He Wang, Dengrong Jiang, Peng Hu, Fuhua Yan, Hao Li, Xingfeng Shao

## Abstract

Accurate quantification of arterial blood T_2_ can be useful for non-invasive assessment of blood oxygenation and blood-brain barrier (BBB) function. While arterial spin labeling (ASL) combined with multi-echo readouts offers a contrast-agent-free approach to map arterial blood T_2_, in vivo applications remain challenging due to rapid signal decay and low signal-to-noise ratio (SNR) at longer echo times (TEs), likely leading to overestimation of T_2_ values. We propose a novel temporal evolution acquisition based ASL (TEA-ASL) sequence incorporating an optimized variable refocusing flip angle (RFA) train to preserve signal across all TEs. Data were acquired on a 5T MRI system combining a pseudo-continuous ASL (pCASL) with the proposed TEA readout with 12 echo times (32-384 ms). The variable RFA scheme significantly improved signal stability across the echo train compared to conventional acquisition with constant RFAs. Accuracy and clinical feasibility of the proposed method was validated by simulations, phantom scans, in-vivo test/retest experiments and in a patient with middle cerebral artery stenosis. The proposed TEA-ASL technique provides robust arterial T_2_ mapping at ultra-high field, offering a promising tool for probing oxygenation-related hemodynamics and BBB-associated pathophysiology.

## 1. Introduction

The blood-brain barrier (BBB) is a highly selective interface formed by cerebral endothelial cells sealed with tight junctions, which critically regulates the exchange of oxygen, nutrients, metabolites, and pathogens between the bloodstream and the central nervous system[1–4]. Both animal and human studies have proved a natural decline or breakdown of BBB while ageing[5, 6]. Growing evidence further links BBB dysfunction to a spectrum of neurological disorders including multiple sclerosis, stroke, traumatic brain injury, Parkinson’s Disease(PD), cerebral small vessel disease (cSVD) and Alzheimer’s Disease (AD)[7–16].

Current clinical approaches to assess BBB permeability rely heavily on exogenous contrast agents, such as gadolinium-based compounds in dynamic contrast-enhanced (DCE) MRI or radiotracers in positron emission tomography (PET)[17]. These techniques have risks including nephrogenic systemic fibrosis, gadolinium deposition in the brain, and radiation exposure[18–22]. And they are often insensitive to mild or early-stage barrier disruption due to the relatively large molecular size of contrast agents[23]. However, the BBB also restricts the passive diffusion of water molecules, which primarily cross via aquaporin-4 (AQP-4) water channels[24, 25]. This limited water permeability provides a unique physiological basis for using water exchange rate across the BBB as a non-invasive biomarker of barrier integrity, particularly in early or subtle pathological states where conventional contrast-based methods may lack sensitivity. ASL MRI is a non-invasive MRI method that magnetically labels arterial water as an intrinsic tracer to quantify cerebral perfusion[26]. Recent advances have extended ASL to probe BBB water permeability by resolving the dynamics of labeled water as it transitions from intravascular to extravascular compartments[27–30], in which a key parameter is the transverse decay of the perfusion signal i.e., the arterial blood T_2_ reflecting blood oxygenation and critically modulates the signal decay of labeled water during its transit across the BBB.

Recently, non-invasive MRI methods combining ASL with T_2_-sensitive readout, such as multi-echo gradient and spin echo (GRASE) sequences or T_2_-preparation modules, have been proposed to map arterial blood T_2_ without exogenous contrast agents[31–33]. There is growing recognition that accurate measurement of arterial T_2_ is essential for advancing non-invasive BBB assessment in neurodegenerative orders like AD and even subtle changes during ageing and caffeine intake, as demonstrated in pre-clinical studies and in vivo studies[34–37]. Accurate estimation of arterial T_2_ enables differentiation between capillary and tissue contributions in ASL signals, thereby facilitating quantification of water exchange rate. However, conventional implementations face significant challenges: a traditional GRASE samples each k-space partition (slice) after each refocusing pulse during the first two TRs (interleaved from center to outer k-space[31]), and rapid signal decay at longer TEs will lead to slice blurring in reconstructed 3D images. Also, the inherently low SNR at longer TEs might lead to overestimation of arterial blood T_2_ so that a long acquisition (∼10 mins) with more averages is typically needed to improve reliability[38].

Furthermore, ASL at UHF gains SNR due to superlinear SNR increase with field strength and longer arterial blood T_1_ [39, 40], and ∼2.42/8.03 fold SNR increase can be achieved when acquiring ASL at 5T as compared to 3T/1.5T[41, 42]. Thus, acquiring ASL at UHF offers a unique opportunity to extend the echo train and capture the full T_2_ decay curve with improved robustness to noise at 5T. Additionally, ∼60-70% *B*^+^ can be achieved at the labeling plane (typically below cerebellum) with a 2Tx head coil, which mitigates the SNR loss due to compromised labeling efficiency at UHF systems.

The purpose of this work is to investigate the feasibility and accuracy of a novel TEA-ASL sequence, incorporating an optimized RFA train, for mapping arterial blood T_2_ in the human brain at 5T. We conducted comprehensive numerical simulations, phantom validation and in vivo experiments in a cohort of healthy participants and one patient with ischemia to evaluate how this approach mitigates signal decay and improves the precision of arterial blood T_2_ mapping.

## 2. Methods

### 2.1 Data Acquisition

All data were acquired on a United Imaging uMR Jupiter 5T MRI system (United Imaging Healthcare, Shanghai, China) with a 2Tx/48Rx head coil. In this study, two protocols were used: the protocol 1 (henceforth referred to the proposed “variable RFA TEA-ASL” protocol) aimed to enable accurate quantification of arterial blood T_2_ by improving signal stability at longer TEs through a variable RFA scheme. To evaluate the reproducibility of the measured arterial blood T_2_, the same protocol was repeated on a separate day in a six-participant subset (1 Female, age: 28.3±3.9 years). The protocol 2 (henceforth referred to as the “constant RFA TEA-ASL” protocol) was also acquired for comparison, acquired side by side in another independent subset of participants. Twelve healthy volunteers (4 Female, age: 24.5±3.8 years) and one patient with right MCA stenosis (Male, 55∼60 years) were recruited in this study. All participants were provided written informed consent in accordance with our institution’s IRB regulations (IRB number: RJ2025300).

The proposed protocol 1 (sequence diagram shown in Fig. 1) consisted of one pCASL scan with labeling duration (LD) = 1800 ms and post-labeling delay (PLD) = 1500 ms, and an M_0_ scan was acquired for calibration. A 3D GRASE readout was employed with TEA so that full kspaces can be acquired across all TEs: the order of partition (slice) encoding is shifted between TRs, so that interleaved kz line at each echo will be sampled by different TRs. By retrospectively recombining the k-space acquired across all TRs (bottom row), fully sampled k-space at each TE can be obtained. The proposed TEA echo train incorporates a variable refocusing angle scheme optimized to preserve signal at later echoes. The readout was segmented into 12 separate acquisitions, in which signal from one echo was collected, along the partition-encoding direction. The values of variable RFAs were optimized in the range of α ∈ [100:20:180] degrees to maximize the sum of signal collected at the last three echoes (echo 9∼12) while keeping the signal generally decreasing except one turning point at each echo for its stability against noise. The signal was simulated using the extended phase graph (EPG) approach[43].

**Figure 1.**
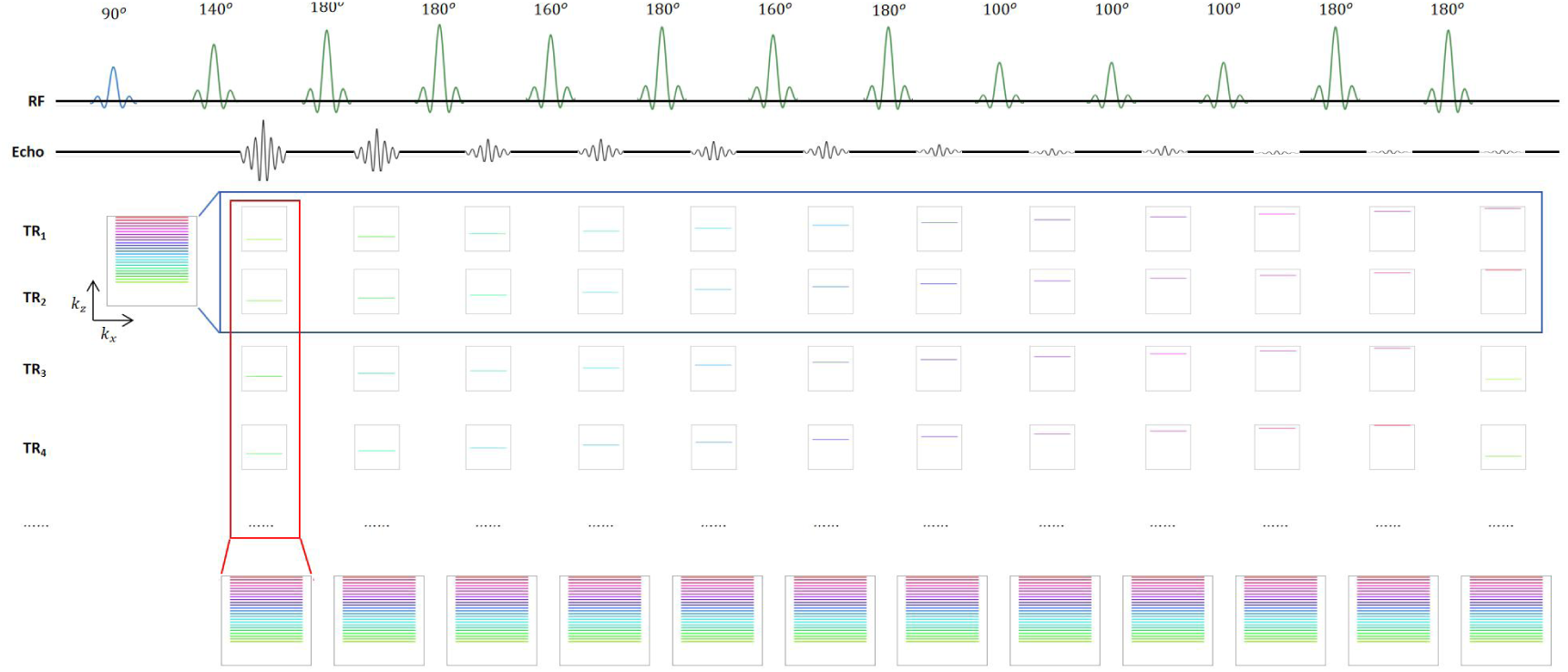
Illustration of the temporal evolution acquisition (TEA) scheme. In this example (32 slices in total and 24 slices collected), traditional GRASE samples each k-space partition (slice) after each refocusing pulse during the first two TRs (interleaved from center to outer k-space, each k-z line is indicated by a different color), and signal decay at longer TEs will lead to slice blurring in reconstructed 3D images. In TEA, the order of partition (slice) encoding is shifted between TRs, so that interleaved k-z line at each echo will be sampled by different TRs. By retrospectively recombining the k-space acquired across all TRs (bottom row), fully sampled k-space at each TE can be obtained.

Imaging parameters were: matrix size = 64×64, 32 slices with 3/4 partial Fourier acquisition, spatial resolution = 3.5 mm³ isotropic, two hyperbolic secant (HS) pulses were utilized for background suppression, TE = ń32 ms (n = 1-12), TR = 6070 ms, acquisition time = 4 min 45 sec.

The protocol 2 shared the same TEA-ASL acquisition as protocol 1, while the echo train used a train of constant RFAs of 180 degrees. The constant RFAs were commonly used in conventional multi-echo GRASE acquisitions. Perfusion signals acquired with protocol 2 were compared to the proposed protocol 1 to demonstrate that perfusion signals can be preserved at later TEs to improve the accuracy of arterial blood T_2_ mapping.

### 2.2 Pre-processing

A brain mask was created to remove any signal from the background by thresholding the average M_0_ image. ASL control and label images were then pair-wise subtracted to generate perfusion-weighted images at all TEs. The resulting multi-echo perfusion signals (N = 12 echoes) from the proposed protocol 1 were subsequently subjected to voxel-wise EPG dictionary matching to estimate the T_2_ of arterial blood.

### 2.3 Voxel-wise Analysis

The ASL signal acquired with protocol 2 follows a mono-exponential decay S = S_0_·e^-TE/T2^, where S_0_ denotes the perfusion-weighted signal amplitude at TE = 0 (i.e., the ideal initial signal before any T_2_ decay). However, the signal collected with the proposed protocol 1 can not be modeled by mono-exponential decay due to non-180° refocusing pulses, and we adopted a dictionary-based signal decay modeling approach. We precomputed a comprehensive dictionary **D∈R**^n^ containing simulated signal trajectories across 12 echo times using the EPG approach. Each column ***d****_k_* corresponds to a unique combination of physiological and instrumental parameters: longitudinal relaxation time T_1_, transverse relaxation time T_2_, and transmit field scaling factor B_1_. The parameter space spans physiologically reliable ranges: *T*_1_∈[1000:10:1600] ms, *T*_2_∈[40:1:200] ms, *B*_1_∈[0.8:0.1:1.2]. This grid yields 49,105 atoms.

For each voxel, the measured multi-echo ASL signal **S∈R***^N^* (where *N* is the number of echoes and N = 12 here) was modeled as:

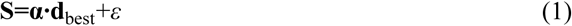

where **d**_best_ is the best-matching atom from **D**, *α* is scaling factor, and *ε* is the residual error accounting for noise, model mismatch, and unmodeled physiological fluctuations.

The matching procedure was performed as follows: For each dictionary atom ***d****_k_*(column *k* of **D**), the optimal scaling factor was computed via orthogonal projection:

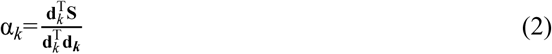

This is the closed-form solution to the least-squares problem min_a_||ad_k_-S||^2^_2_. The corresponding fitted signal was then **S^^^**=*α_k_***d***_k_*, and the residual error was quantified as:

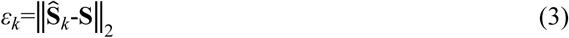

The atom yielding the minimum residual error was selected as the best match:

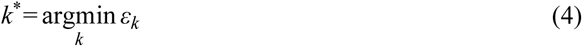

Thus the effective signal decay pattern was defined by **d*_k_\****. Since each atom is associated with a known T_1_, T_2_, B_1_ triplet, this selection implicitly provides estimates of these underlying parameters. In practice, we first quantified the local *B*^+^ effect and generate a *B*_1_ map using control signals with higher SNR. Subsequently, we selected the optimal atom matching the decay pattern of the perfusion signal under the voxel-specific *B*^+^ value derived from the *B*^+^ map, using equations (2)–(4).

### 2.4 Total Generalized Variation (TGV) Based Denoising

TGV-based algorithm has been used for reconstruction of undersampled ASL signal[44], in which the unknown image *u* is estimated from noisy measurements *i* by solving a regularized optimization problem, generally expressed as:

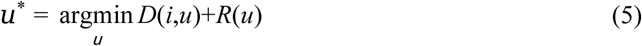

where *D(i, u)* denotes the data-fidelity term that links the fitted data *u* to the collected image *i* and *R(u)* denotes the regularization term.

In this work, we applied TGV as a post-processing denoising strategy to improve the reliability of T_2_ maps. Specifically, after voxel-wise dictionary-based matching of the multi-echo perfusion-weighted signals, the resulting T_2_ parameter map was often corrupted by noise. To address this issue, we formulated a regularization-based optimization problem that directly acted on the estimated T_2_ map (denoted as *u*), rather than on the image time series or k-space data.

The denoising problem was cast in a general variational form same as Eq. (5) and here *D(i,u)* represented a data-fidelity term that enforced closeness between the T_2_ map *u* and the noisy original image *i* through a reverse process of the dictionary matching.

For regularization term *R*, we adopted second-order TGV[45], which had been shown to effectively preserve edges. TGV was particularly well-suited for T_2_ maps, which were expected to be voxel-wise smooth across anatomical regions. The TGV penalty was defined as:

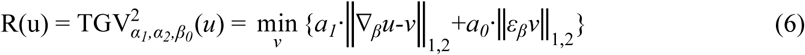

where ∇ *u*=(*β_1_ δ_x_ u,β_1_ δ_x_ u,β_1_ δ_y_ u,β_2_ δ_1_ u*) and *εv* = ½(∇_β_*v*+(∇_β_*v*)^T^) denoted the gradient and symmetrized gradient operators, respectively. The final denoising problem thus read:

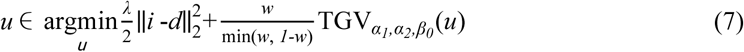

where *u*∈R*^Nx×Ny×Nz^*was the denoised T_2_ map and *d*∈R*^Nx×Ny×Nz×N^*was composed of the **α·d**_best_ value in Eq.1 corresponding to each point in the T_2_ map and the corresponding T_1_ and B_1_ values fitted from the control signal Figure 2 compares the estimated T_2_ values before and after TGV denoising, demonstrating substantial suppression of voxel-wise noise with preservation of the contrast among GM, WM and CSF.

**Figure 2.**
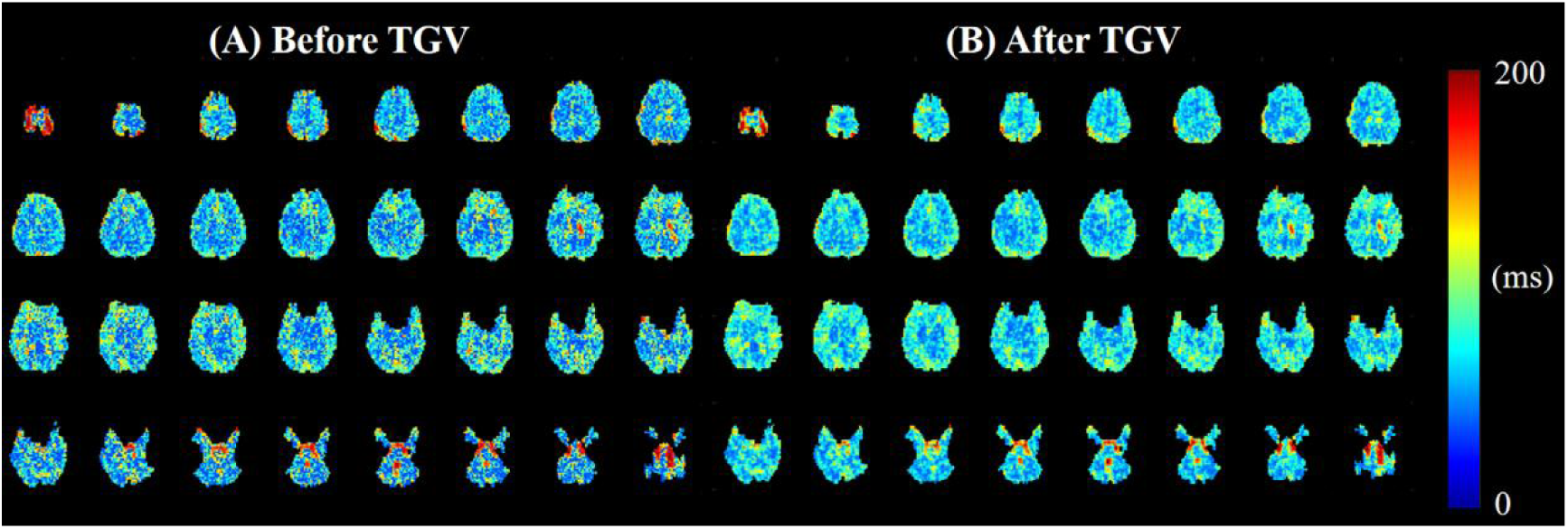
Comparison of estimated T2 values before (A) and after (B) TGV denoising.

### 2.5 Numerical Simulation

To evaluate the robustness and accuracy of the proposed dictionary-based fitting approach in comparison to conventional mono-exponential modeling, a series of controlled numerical simulations were conducted across a range of SNR levels. Two representative SNR conditions were selected to reflect realistic scenarios encountered in ASL experiments at ultra-high field: SNR = 5 and 30, for simulating low and high perfusion signals.

For each SNR level, 100 independent simulated signals with noise were generated for both the proposed dictionary matching method and a standard mono-exponential model. To enable a fair comparison, comprehensive signal dictionaries were precomputed separately for each of the two protocols under identical modeling conditions The underlying noise-free signal was synthesized using EPG formalism to accurately model the magnetization evolution under the specific TEA-ASL readout scheme. The EPG simulation incorporated realistic sequence timing (echo spacing = 32 ms), and the resulting signal amplitudes at each echo time were computed for a range of physiological parameter combinations ( *T*_1_∈ [1000:10:1600] ms, *T*_2_∈ [40:1:200] ms, *B*_1_∈[0.8:0.1:1.2]), covering the expected variability in labeled arterial blood and brain tissue at 5T. Complex Gaussian noise with zero mean and standard deviation σ= S/SNR was added to the magnitude of the simulated signal to produce realistic noisy decay curves. Each noisy decay curve was matched to the dictionary by the same method as real signal.

All simulations and fitting procedures were implemented in MATLAB (R2024a, MathWorks Inc), with random number seeds fixed for each noise realization to ensure consistency across methods.

### 2.6 Phantom Validation

To assess the fidelity of the EPG-based dictionary matching framework in estimating relaxation parameters, a commercial MRI phantom (CaliberMRI, Boulder, CO) was scanned using the variable and constant RFA TEA-ASL sequences employed in the in vivo experiments. The phantom comprises an array of spheroidal vials filled with aqueous solutions doped with variable concentrations of paramagnetic contrast agents (manganese chloride in T_2_ calibrated vials, specifically), each formulated to yield distinct and stable T_2_ relaxation times. The manufacturer provides reference T_2_ values for each vial measured at 1.5T and 3T, however, these values are not directly applicable at 5T. We acquired a separate T_2_ map of the phantom using multi-echo GRE sequence as reference (spatial resolution = 0.8×0.8×5 mm^3^, TE = 12.5 ms, TR = 2348ms). The proposed protocol 1 was used to scan the phantom, with 2, 4 and 6 averages to evaluate the quantification accuracy across SNR levels. ROIs corresponding to individual vials were defined manually on the first-echo images using ITK-SNAP (version 4.2.0-rc.1, open-source software; https://itk.org/).

## 3. Results

### 3.1 SNR Preservation at Long TEs through RFA Optimization

The optimized pattern of variable RFAs contains a train of RFAs valued 140°, 180°, 180°, 160°, 180°, 160°, 180°, 100°, 100°, 100°, 180° and 180° in order. Figure 3 shows the comparison of simulated data and collected signal. Compared to the conventional protocol with a train of constant RFAs, the proposed variable RFA sequence enhanced the signal amplitude at the 5∼12 echoes by 58.3% in simulation with physiological reasonable parameters and 39.7% on average in the data collected.

**Figure 3.**
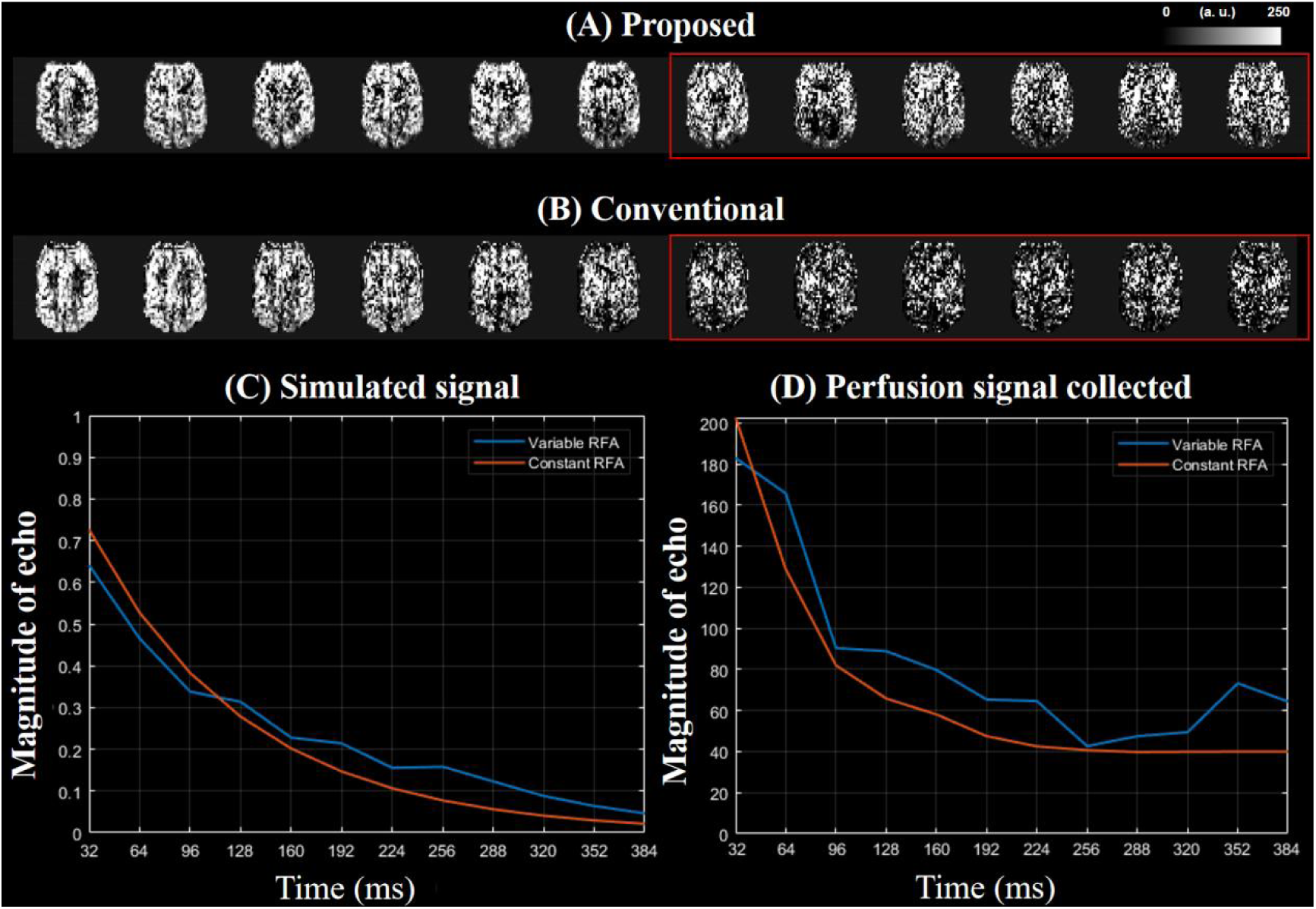
(A) Perfusion signal (label-control) collected with the proposed method. (B) Perfusion signal collected with the conventional constant RFA method. (C) Comparison of simulated signal with EPG for proposed variable RFA method and constant RFA method (D) Comparison of perfusion signal collected with proposed variable RFA method and constant RFA method.

### 3.2 Numerical Simulation

Figure 4 shows simulation results. For low SNR (SNR = 5) data, the protocol 2 exhibited substantial bias and variance in estimating T_2_. In contrast, the proposed protocol 1 demonstrated improved reliability, with 71.6% lower root-mean-square error (RMSE) in T_2_ estimation across all 100 independent experiments. For variable RFA protocol (protocol 1), the estimated T_2_ was 118.5±52.1 ms compared to a reference T_2_ of 114.3±48.3 ms (RMSE = 25.4) while the T_2_ estimated by conventional protocol 2 was 57% overestimated, as expected (138.3±102.9 ms, RMSE = 89.5).

**Figure 4.**
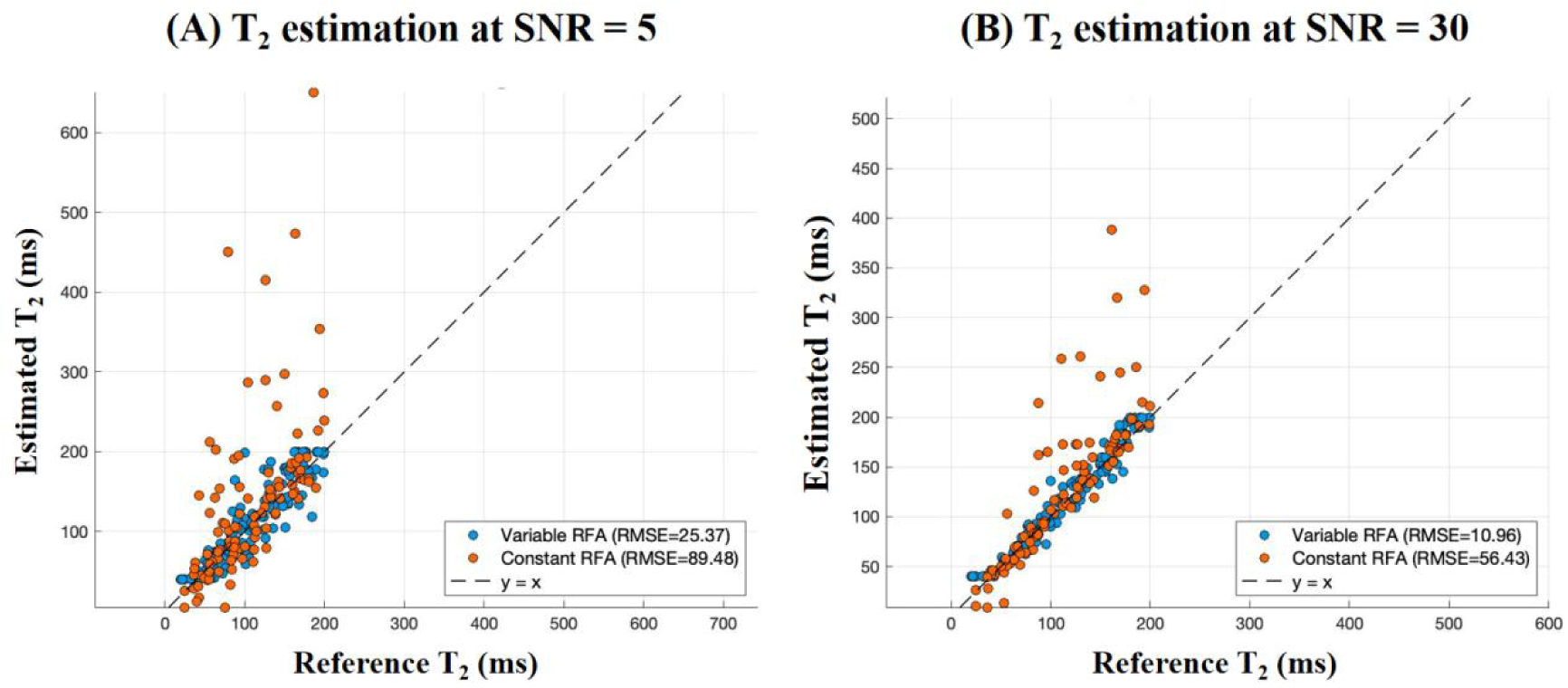
Comparison of estimated T2 values versus reference T2 using the simulated signal of the proposed acquisition with variable RFA and the simulated signal of the conventional multi-echo acquisition with constant RFA when SNR = 5 (A) and 30 (B), respectively.

When simulating high SNR data (SNR=30), protocol 2 showed reduced but still noticeable overestimation and variability in T_2_ estimation (126.1±82.9 ms, RMSE = 56.4). In contrast, the proposed variable RFA strategy (protocol 1) maintained better accuracy and lower RMSE in T_2_ estimates (116.8±48.3 ms, RMSE = 25.4). This advantage stemmed from its ability to retain stable signals at longer TEs.

### 3.3 Validation via Calibrated Phantom Scans

Figure 5 shows reference and estimated T_2_ maps of the T_2_ quantification phantom. Results demonstrated strong agreement between the T_2_ values estimated by the proposed EPG-based dictionary approach and those obtained from the product T_2_ mapping sequence as reference. Across all vials and SNR levels (NEX = 2, 4, 6), the proposed dictionary-derived T_2_ estimates consistently tracked the reference values with minimal deviation, exhibiting both high accuracy and reproducibility (RMSE = 24.1, 25.5 and 14.1 for NEX = 2, 4 and 6, respectively. Details listed in Table S1, same for conventional fit). In contrast, the mono-exponential fit exhibited biased estimation of T_2_, particularly in vials with extra-long relaxation times (> 200 ms), where the discrepancy between ideal exponential decay and the actual signal evolution was most apparent (RMSE = 34.0, 33.3 and 8.0 for NEX = 2, 4 and 6, respectively).

**Figure 5.**
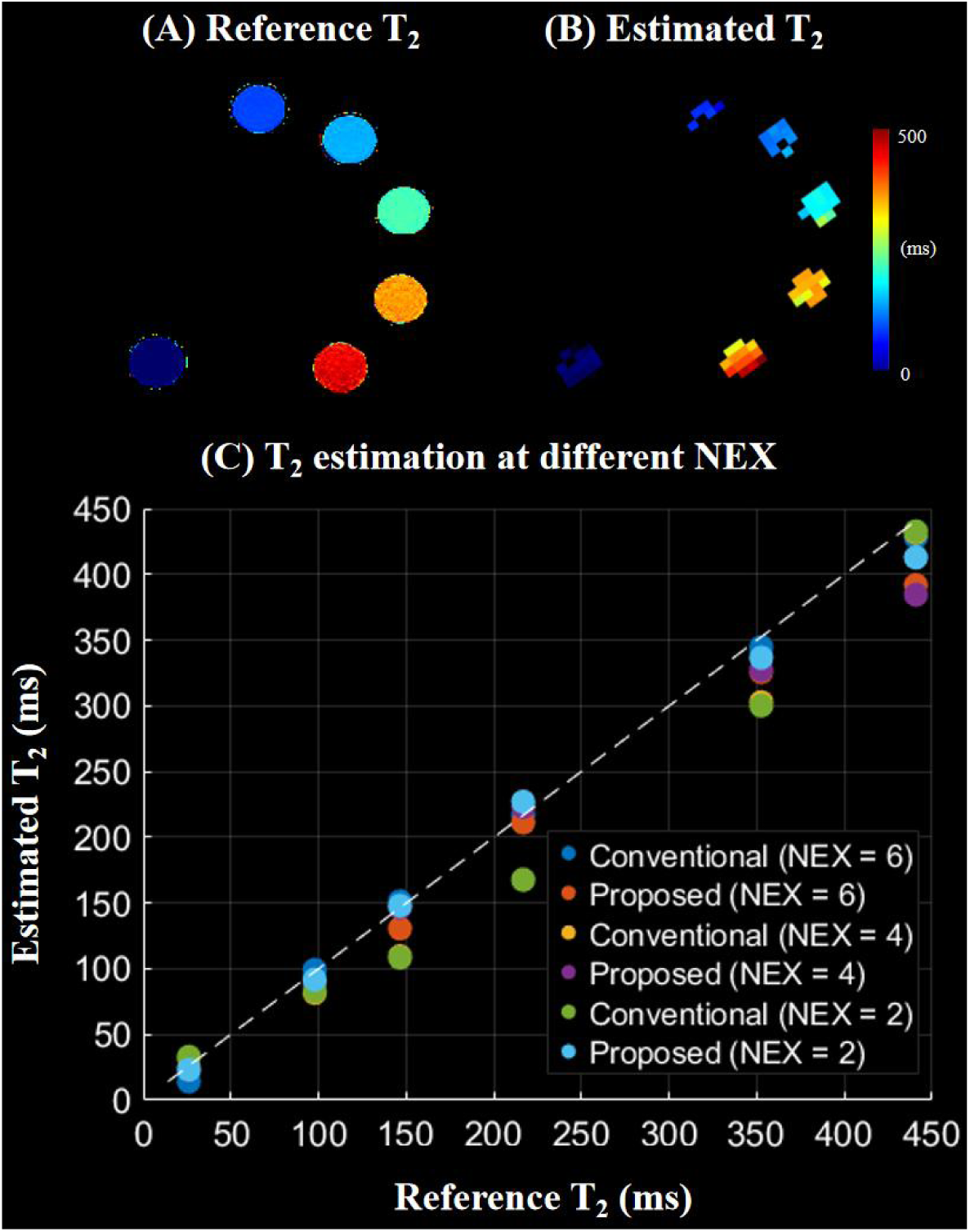
(A) Reference T2 values measured with the built-in product multi-echo GRE T2 mapping sequence. (B) Fitted T2 maps with using the proposed method. (C) Scatter plots of reference and estimated T2 values of the phantom.

### 3.4 In Vivo Experiments

Figure 6 shows estimated T_2_ maps from test and retest scans of a healthy volunteer using the proposed protocol 1 (B and C), and in comparison with results acquired using protocol 2 (A). Voxel-wise fitting of multi-echo perfusion signals acquired with a variable RFA sequence has demonstrated robust and reliable estimation of T_2_ maps in both GM and WM between test and retest scans, exhibiting high spatial consistency and physiological plausibility. The average arterial T_2_ value was 64.8±3.2 ms in gray matter and 66.4±3.1 ms in white matter (slightly longer in WM due to prolonged ATT and likely higher oxygenation level[46].

**Figure 6.**
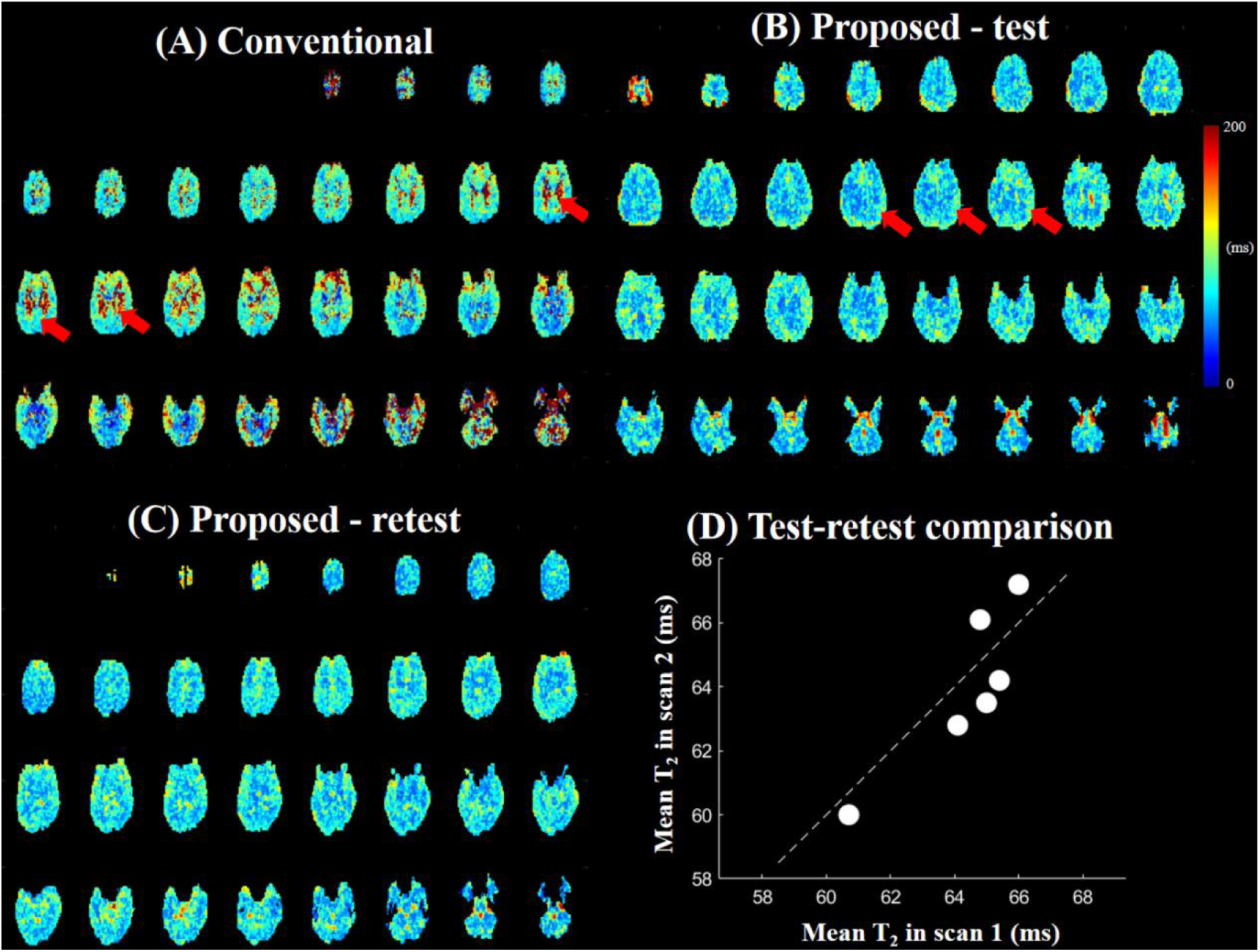
Fitting result with the signal of a healthy volunteer collected through (A) a constant RFA protocol and (B, C) The proposed variable RFA protocol. (D) Scatter plots of whole brain averaged arterial blood T2 between test and retest scans(ICC=0.884).

In contrast, a systematic overestimation of T_2_ (protocol 2) can be observed in Figure 6 (A), particularly in WM. This bias arises primarily from the inherently lower SNR of ASL signals in WM (∼1/3) compared to GM. In conventional spin-echo trains with constant 180° refocusing pulses, the signal from WM decays rapidly due to its shorter intrinsic T_2_, and by the fifth echo (TE = 160 ms), the magnitude often approaches the noise floor. At later echo times, where the signal from WM has already decayed, the low SNR leads to increased uncertainty in the measured signals. When fitting T_2_ values from multi-echo signals, this noise-induced variability can cause the algorithm to misattribute random fluctuations as slower decay, thereby resulting in overestimation of T_2_ values.

In a patient with right MCA stenosis (Figure 7), significantly elevated arterial T_2_ in the affected region (137.4 ± 11.6 ms vs 66.8 ± 17.4 ms at the lesion and contralateral side) was observed (Figure 7 (A)). This result is likely linked to delayed flow and higher oxygenation level due to prolonged transit, which agreed well with the ATT result (Figure 7 (B)), further validating the accuracy of the T_2_ mapping using the proposed method.

**Figure 7.**
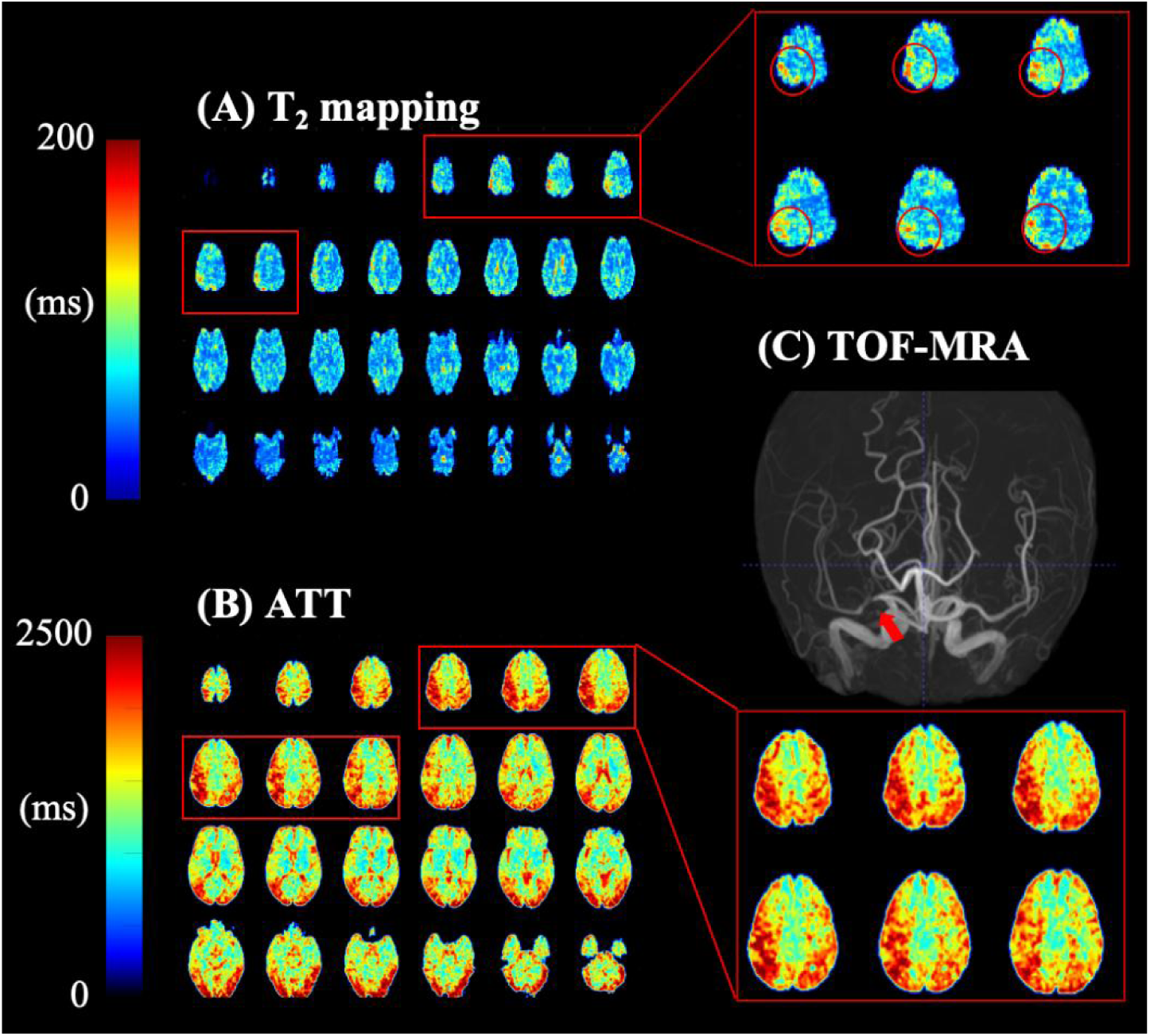
Estimated T2 maps (A) of a patient with right MCA stenosis (Regions with increased T2 values likely induced by delayed flow was marked by red circles). For comparison, ATT maps calculated from multi-delay ASL was also shown in B. (C) Time of flight (TOF) result of the corresponding patient with stenosis location indicated by a red arrow.

## 4. Discussion

In this study, we introduced TEA-ASL acquisition that reorganizes the temporal-spatial relationship between echo time and k-space sampling. Rather than assigning fixed slice encodings to specific echoes within a single TR, TEA-ASL distributes partition encoding across multiple TRs such that each echo time ultimately samples a fully interleaved, retrospectively reconstructable k-space. This decoupling of T_2_ decay from spatial encoding ensures that we can conduct robust voxel-wise T_2_ fitting without the confounding influence of slice-dependent point spread function degradation. To address the rapid signal decay that plagues multi-echo ASL signals at long TEs, we implemented a variable RFA scheme optimized via EPG simulations, which accounts for a slower decay in the middle of the echo train, to maximize signal retention. These strategies make possible an accurate quantification of arterial blood T_2_, which can be useful for probing cerebral blood oxygenation since deoxygenated hemoglobin is paramagnetic and significantly shortens blood T_2_, whereas oxygen-rich blood exhibits a longer T_2_ value[47]. And arterial blood T_2_ has also been incorporated with emerging models of BBB water permeability/exchange measurements by distinguishing intravascular and extravascular compartments[34].

To estimate arterial blood T_2_ from TEA-ASL signals, we adopted an EPG-based dictionary matching framework. Since there is not an analytical solution to the signal derived from variable RFA, we precomputed a comprehensive dictionary of simulated signal evolutions across a physiologically plausible parameter space[48]. A second-order TGV-based approach was further utilized to improve spatial smoothness while maintaining signal fidelity given the low SNR of ASL signal[38].

In simulation and phantom studies, the variable RFA strategy significantly improved the robustness of T_2_ estimation compared to conventional constant RFA. Under low-SNR conditions, constant RFA led to biased and unstable T_2_ estimates, especially for longer T_2_ values, because the fitting relied heavily on later echoes that were dominated by noise. In contrast, the proposed variable RFA scheme was designed to preserve ASL signals at longer TEs. With sufficient signal acquired even at late echoes, this approach provided additional temporal information that helped disentangle true T_2_ values from random noise fluctuations, and our simulation and phantom results demonstrated that the proposed method can provide accurate T_2_ mapping at different SNR levels.

In vivo experiments in healthy volunteers further validated findings from simulation and phantom studies. The proposed TEA-ASL can generate reliable voxel-wise T_2_ mapping in both gray and white matter, yielding spatially consistent results with high test and retest reproducibility (ICC = 0.884). Interestingly, we found markablly high T_2_ values in ventricles and choroid plexus, this suggests that the labeled protons in the blood entered CSF, further revealing the potential of the proposed methods for quantification blood-CSF exchange. In contrast, results from protocol 2 generated systematical higher T_2_ values, particularly in white matter, where intrinsic SNR is lower and signal decays more rapidly across TEs. We hypothesized that the WM signals often dropped to near the noise floor by mid-to-late echoes (by the fifth echo i.e. TE = 160 ms in our experiment). Fitting algorithms then misinterpreted noise as slower decay, leading to overestimated T_2_. By sustaining sufficient TEA-ASL signals at later TEs, the proposed protocol 1 makes it suited for robust and accurate quantitative in-vivo T_2_ mapping.

Our observation of physiologically reliable and reasonable T_2_ values together with elevated T_2_ in an MCA stenosis patient demonstrated the method’s sensitive and robust measurement of the apparent T_2_ that depends on contribution of the signal from the IV and EV space as described above. Therefore, our proposed method can potentially be further used for BBB assessment with a sensitive and robust measurement of the relative contributions of the IV space and EV tissue within the overall ASL signal in the future.

When neurovascular unit function is compromised, it can impair the efficacy of the delivery of oxygen and essential nutrients to brain tissue, leading to bioenergetic deficits that jeopardize neuronal viability. Concurrently, BBB disruption may permit harmful, blood-derived neurotoxins to infiltrate the brain parenchyma. At earlier stages, dysfunction of the glymphatic system may further exacerbate this toxic milieu by reducing clearance of endogenous metabolic waste, such as beta-amyloid peptides[49]. Together, these interrelated pathophysiological processes can drive chronic neuroinflammation, progressively erode neuronal health, and ultimately contribute to neurodegeneration, as observed in AD and normal ageing[1, 50–54].

Notably, growing evidence suggests that BBB breakdown may serve as an early initiating event in both aging and AD pathogenesis, and it is increasingly implicated across a broad spectrum of neurological disorders[55]. Given that arterial T_2_ measured by TEA-ASL reflects the relative contributions of IV and EV signal—and is thus sensitive to changes in BBB permeability and microvascular integrity—future studies could leverage this technique to detect early vascular dysfunction, stratify disease severity, or map regional heterogeneity in arterial T_2_ as a noninvasive biomarker of neurovascular health.

As mentioned above, the TEA-ASL at 5T gain an ∼2.42-fold SNR gain compared to 3 T, which allows acquiring higher spatial resolution (3.5 mm^3^ isotropic), as compared to recent 3 T protocols of 3.5×3.5×8 mm^3^ for BBB water exchange mapping [36, 56]. Moreover, our sequence utilized 12 echoes, extending beyond the 8 echoes or less typically feasible at 3T[37]. This extended echo train is critical because later echoes, which are essential for accurate T_2_ estimation, are often buried in noise at lower field strengths, where SNR constraints severely limit reliable sampling of the signal decay curve.

There are several limitations of this study. Firstly, the current segmented acquisition introduces vulnerability to motion-induced phase inconsistencies between segments. Although k-space motion correction was applied, residual artifacts may persist in patients with severe motion. Secondly, current spatial resolution is 3.5 mm^3^ isotropic, and it remains challenging to study small brain regions such as subcortical nuclei (e.g., thalamus, basal ganglia). Future work could explore accelerated strategies like CAIPIRINHA acquisition or AI-based methods to reduce motion sensitivity while enhancing the overall SNR and resolution[57–59]. Thirdly, the current quantification model assumes a single T_2_ value within one voxel, and potential multi-component relaxation (e.g., intra- and extracellular water pools) effects will be studied in the future incorporating multi-pool EPG models[60]. On top of that, it is necessary to carry out clinical validation involving more diverse disease indications and a greater patient cohort.

## 5. Conclusion

In conclusion, the proposed TEA-ASL with variable RFA, EPG modeling, and TGV denoising represents a promising method for accurate arterial blood T_2_ mapping at 5T, which offers strong potential as a surrogate biomarker for non-invasive assessment of microvascular and BBB function.

## Data Availability

Data and code used in this paper can be shared upon request to the corresponding author.

