## Supplementary material for "Temporal evolution acquisition based arterial spin labeling (TEA-ASL) for accurate arterial blood T_2_ mapping": Table S1

Table S1. Summary of different spheres under different scans. All values are shown as mean±SD

|  | Reference | Conventional  (NEX=6) | Proposed  (NEX=6) | Conventional  (NEX=4) | Proposed  (NEX=4) | Conventional  (NEX=2) | Proposed  (NEX=2) |
| --- | --- | --- | --- | --- | --- | --- | --- |
| Sphere #1 | 440.7±31.9 | 428.3±15.4 | 412.8±90.7 | 431.7±24.0 | 384.6±52.8 | 432.7±25.5 | 391.8±57.0 |
| Sphere #2 | 352.5±24.1 | 344.6±21.7 | 336.6±64.0 | 302.8±25.3 | 327.1±55.4 | 300.2±25.8 | 325.7±55.2 |
| Sphere #3 | 216.7±10.8 | 216.7±14.8 | 227.1±62.4 | 167.8±14.6 | 223.1±58.9 | 167.6±14.5 | 211.2±48.6 |
| Sphere #4 | 146.5±15.3 | 151.3±15.4 | 147.9±28.8 | 109.4±8.5 | 146.8±29.0 | 108.3±8.4 | 130.6±12.1 |
| Sphere #5 | 97.8±30.4 | 99.2±29.5 | 91.7±19.5 | 81.6±20.9 | 89.8±17.2 | 82.8±23.3 | 88.9±19.5 |
| Sphere #9 | 26.1±2.0 | 14.2±5.0 | 23.1±8.7 | 32.6±5.7 | 23.0±8.6 | 33.0±6.1 | 24.2±7.7 |
| RMSE |  | 8.0 | 14.1 | 33.3 | 25.6 | 34.0 | 24.1 |
