## Supplementary material for "Temporal evolution acquisition based arterial spin labeling (TEA-ASL) for accurate arterial blood T_2_ mapping": Table S2

Table S2. Summary of estimated T_2_ value in WM and GM of all healthy volunteers. All values are shown as mean±SD

| Subject # (sex) | GM | WM |
| --- | --- | --- |
| 1 (M) | 63.9±4.7 | 66.9±6.3 |
| 2 (M) | 60.9±4.2 | 68.7±6.3 |
| 3 (M) | 65.1±4.5 | 64.9±7.0 |
| 4 (M) | 65.3±5.1 | 62.9±6.9 |
| 5 (F) | 65.9±4.6 | 66.1±6.5 |
| 6 (F) | 58.5±4.6 | 62.9±4.5 |
| 7 (M) | 64.9±6.8 | 72.9±8.0 |
| 8 (F) | 71.5±7.4 | 63.4±5.2 |
| 9 (M) | 65.1±4.0 | 68.8±8.5 |
| 10 (F) | 65.5±7.5 | 63.8±8.4 |
| 11 (M) | 68.0±6.0 | 69.4±9.3 |
| 12 (M) | 63.1±8.1 | 65.7±8.2 |
| Mean (4F8M, 25.8±4.5) | 64.8±3.2 | 66.4±3.1 |
